# Early Fc-effector antibody signatures impact COVID-19 disease trajectory

**DOI:** 10.64898/2026.02.18.26346542

**Authors:** Alba Escalera, Ana S. Gonzalez-Reiche, Sadaf Aslam, Enrique Bernal, Galit Alter, Amaya Rojo-Fernandez, Alexander Rombauts, Gabriela Abelenda-Alonso, Mary Anne Amper, Venugopalan D. Nair, Harm van Bakel, Jordi Carratalà, Adolfo García-Sastre, Teresa Aydillo

## Abstract

Why do some individuals develop mild COVID-19 while others progress to severe disease remains a central challenge in SARS-CoV-2 immunology. In this study, we leveraged the BACO Cohort – a unique historical cohort of immunologically naïve, hospitalized COVID-19 patients from the first pandemic wave – to investigate early immune determinants of clinical disease trajectories. Integrating bulk RNA-seq, Olink proteomics, and systems serology, we identified two fundamentally distinct immune trajectories according to disease phenotypes. Severe patients exhibited upregulation of proinflammatory genes and monocyte-associated transcripts, alongside downregulation of genes related to T cell responses and immune signaling. Notably, an upregulation of inhibitory Fc-receptor–associated gene was also found in severe cases. In contrast, mild cases showed coordinated lymphoid activation and limited inflammation. Building on these findings, we performed a functional profiling of Fc-effector activity in the polyclonal serum of the patients and found that monocyte-mediated phagocytosis was a common feature of mild disease. Interestingly, this response was mainly driven by rapid induction of S1-specific antibodies. Conversely, severe patients tended to generate higher levels of S2-biased antibodies early after infection with poor Fc-effector functionality. Together, these findings demonstrate that early S1-directed, Fc-competent humoral immunity is a key determinant of favorable COVID-19 outcomes, while delayed functional maturation and early S2 bias characterized severe disease in the BACO cohort.

## INTRODUCTION

Severe Acute Respiratory Syndrome Coronavirus 2 (SARS-CoV-2) emerged in Wuhan (China) in 2019 causing more than 778 million infections and 7 million deaths worldwide as of December 24, 2025 (https://covid19.who.int/). The clinical manifestations of coronavirus disease 2019 (COVID-19) are highly diverse, ranging from asymptomatic cases to symptomatic presentations. While most infections cause mild symptoms, a subset of cases can progress to severe or critical illness, leading to fatal outcomes^1,2^. Several efforts were made in the early pandemic to understand the underlying immune features associated with patient risk and to identify potential therapeutic targets. Initial studies using both bulk RNA and single-cell sequencing technologies revealed distinct immune signatures according to COVID-19 disease phenotype^3–6^. Patients with mild COVID-19 often presented controlled inflammatory responses and early activation of antiviral pathways, such as type I interferon signaling^7,8^. In contrast, severe patients showed higher viral loads and high levels of inflammatory markers, including C-reactive protein (CRP) and ferritin^7,9–11^. Similar to severe influenza infection^12–14^, disease severity was also characterized by a canonical cytokine storm-like with increased serum levels of IL-1β, IL-6, IL-8, TNFα ^2,7,15–19^. Severe COVID-19 was also associated with marked lymphopenia, T cell exhaustion and increased numbers of inflammatory monocytes^10,17,20,21^. Besides, risk factors such as age and comorbidities have been linked to poor outcomes^22–24^. However, young and healthy individuals can also experience severe illness, underscoring the complexity of the immune response against SARS-CoV-2. Therefore, many questions remain still unanswered regarding the immunological and molecular mechanisms of disease severity.

The B cell compartment constitutes a major defense against viral infections by producing virus-specific antibodies and supporting long-term immune memory. For SARS-CoV-2, antibodies against the spike (S) protein can neutralize the virus and provide protection from infection, especially when targeting the S1 receptor-binding domain (RBD) and N-terminal domain (NTD)^25–27^. However, this neutralization ability is largely strain-specific and can be evaded by virus evolution. In contrast, antibodies against the S2 domain are broadly cross-reactive among human beta-coronaviruses (HCoVs) but have limited neutralization capacity^28–30^. On the other hand, antibodies can also mediate other antiviral functions beyond neutralization through Fc-effector functions. Some of these include antibody-dependent cellular cytotoxicity (ADCC) and antibody-dependent cellular phagocytosis (ADCP). ADCC is primarily mediated by the activating receptor FcγRIIIa and the inhibitory receptor FcγRIIIb, whereas ADCP involves engagement with the activating FcγRIIa and the inhibitory FcγRIIb^31,32^. Interestingly, polymorphisms in FcγR genes can result in allelic variants with distinct functional capacities^33^, some of them still unknown. While Fc-mediated antibody functions have been well characterized in the context of other viral infections, little is known in the case of SARS-CoV-2. Us^34^ and others^35–38^ have previously demonstrated that non-neutralizing antibodies against the conserved hemagglutinin stalk, but not the globular head domain, can mediate Fc-effector functions and confer protection from infection and severe disease. Whether antibodies against SARS-CoV-2 proteins can play a role in COVID-19 outcomes through Fc-activity is not known. In addition, it is still unclear whether SARS-CoV-2 antibodies with Fc-effector functionality could preferentially target the S1 versus S2 domains of the S protein.

At present, most studies aiming to define immune predictors of COVID-19 severity necessarily rely on recent clinical cohorts of individuals with several rounds of SARS-CoV-2 exposure. Either through natural infection or vaccination, this will likely result in heterotypic immunity. This complex immune background makes it difficult to identify the true immune correlates of disease severity during primary infection. In here, we used the BACO Cohort: immunologically naïve COVID-19 patients from the first pandemic wave of SARS-CoV-2 in Spain. The nature of the antibody response in the BACO patients has been partially characterized in our early studies^39,40^. Now, we used this cohort to investigate the mechanisms underlying severe versus mild disease in the absence of SARS-CoV-2 specific memory. First, we performed RNA-seq and proteomic studies. As expected, patients with severe COVID-19 showed impaired T cell activation, enhanced inflammation signatures and monocyte expansion. Interestingly, severe cases also exhibited an early upregulation of inhibitory genes involved in Fc receptor-mediated activity. Next, we applied an integrative functional and systems serology approach. Our data showed that mild patients showed early enhanced Fc-effector ADCP responses, which correlated with the induction of anti-S1 antibodies. Conversely, higher levels of the more conserved and less functionally active anti-S2 antibodies were observed in severe patients at early timepoints. Taken together, our findings provide evidence of the role of anti-S1 Fc-active antibodies in modulating COVID-19 progression. It also underscores the importance of a complete understanding of Fc-mediated functions as determinants of disease trajectory.

## RESULTS

### The BACO Cohort and COVID-19 Disease Severity

The BACO cohort is a unique historical cohort composed of 37 SARS-CoV-2 immunologically naïve, hospitalized patients recruited in Barcelona (Spain) during the first pandemic wave of SARS-CoV-2 (March to May 2020)^39,40^. Mean age of the total cohort was 65 (39-90) years and 25 (67%) of the patients were male. To identify immune determinants of COVID-19 severity, we first stratify the patients according to disease trajectory using a previously published COVID-19 scale ^39,41^. Detailed criteria used for this classification can be found in the Materials and Methods section of this manuscript. Based on this, 26 patients were classified as mild/moderate (referred to as ‘mild’) and 11 as severe/severe end-of organ disease (referred to as ‘severe’). We observed no significant differences in mild versus severe patients in age (64.4 (39-85) vs. 67.9 (47-90) years), sex (male 17 (65.4%) vs. 8 (72.7%) and prevalence of co-morbidities (8 (30.8%) vs. 4 (36.4%). The mean number of days from symptom onset to hospitalization was also comparable among groups (7.3 (3-14) vs. 8.8 (2-14) days). However, symptoms such as throat ache, cough, dyspnea and saturation of oxygen (Sp02<94%) were more frequent in patients with severe disease. Besides, severe patients exhibited an increase in white blood cell and neutrophil counts upon hospital admission. All severe cases and all mild patients, except for one, had SARS-CoV-2 viral pneumonia. Four (36.4%) severe patients required ICU admission and 5 (45.5%) of them died. As expected, mean duration of hospitalization was longer in severe patients (18.4 (3-47) days) compared to those with mild disease (8.2 (2-16) days). Demographics, clinical characteristics, interventions, such as drug therapy, and outcomes are detailed in Table 1.

**Table 1.**
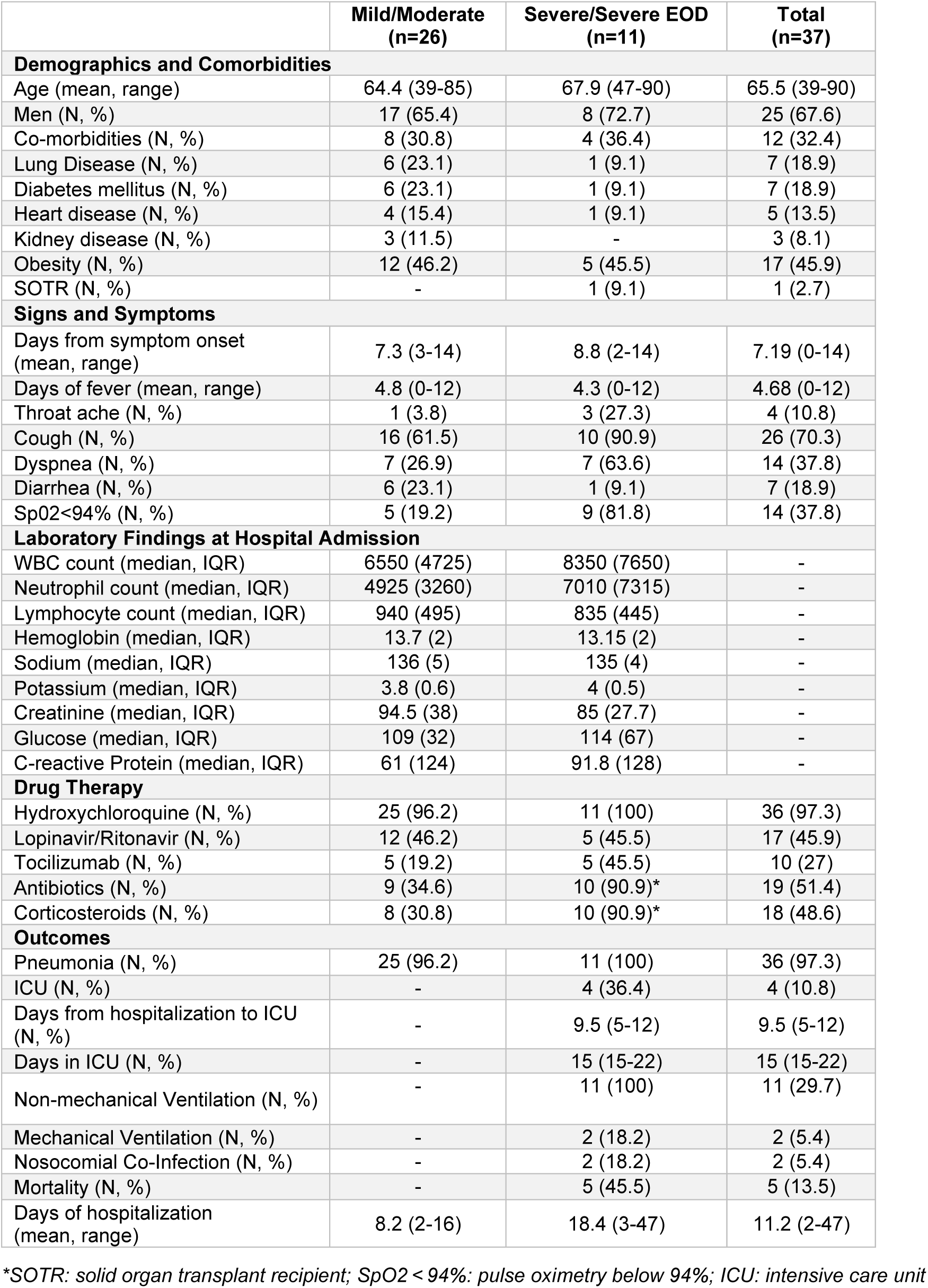
Demographics and clinical characteristics of the BACO cohort according to disease trajectory. Shown are the demographics, comorbidities, signs and symptoms, laboratory findings at hospital admission, drug therapy and outcomes in mild and severe COVID-19 patients from the BACO cohort. An asterisk (*) indicates statistically significant differences (p ≤ 0.05) between groups as determined by the chi-square test.

|  | Mild/Moderate<br>(n=26) | Severe/Severe EOD<br>(n=11) | Total<br>(n=37) |
| --- | --- | --- | --- |
| **Demographics and Comorbidities** |  |  |  |
| Age (mean, range) | 64.4 (39-85) | 67.9 (47-90) | 65.5 (39-90) |
| Men (N, %) | 17 (65.4) | 8 (72.7) | 25 (67.6) |
| Co-morbidities (N, %) | 8 (30.8) | 4 (36.4) | 12 (32.4) |
| Lung Disease (N, %) | 6 (23.1) | 1 (9.1) | 7 (18.9) |
| Diabetes mellitus (N, %) | 6 (23.1) | 1 (9.1) | 7 (18.9) |
| Heart disease (N, %) | 4 (15.4) | 1 (9.1) | 5 (13.5) |
| Kidney disease (N, %) | 3 (11.5) | - | 3 (8.1) |
| Obesity (N, %) | 12 (46.2) | 5 (45.5) | 17 (45.9) |
| SOTR (N, %) | - | 1 (9.1) | 1 (2.7) |
| **Signs and Symptoms** |  |  |  |
| Days from symptom onset (mean, range) | 7.3 (3-14) | 8.8 (2-14) | 7.19 (0-14) |
| Days of fever (mean, range) | 4.8 (0-12) | 4.3 (0-12) | 4.68 (0-12) |
| Throat ache (N, %) | 1 (3.8) | 3 (27.3) | 4 (10.8) |
| Cough (N, %) | 16 (61.5) | 10 (90.9) | 26 (70.3) |
| Dyspnea (N, %) | 7 (26.9) | 7 (63.6) | 14 (37.8) |
| Diarrhea (N, %) | 6 (23.1) | 1 (9.1) | 7 (18.9) |
| SpO2<94% (N, %) | 5 (19.2) | 9 (81.8) | 14 (37.8) |
| **Laboratory Findings at Hospital Admission** |  |  |  |
| WBC count (median, IQR) | 6550 (4725) | 8350 (7650) | - |
| Neutrophil count (median, IQR) | 4925 (3260) | 7010 (7315) | - |
| Lymphocyte count (median, IQR) | 940 (495) | 835 (445) | - |
| Hemoglobin (median, IQR) | 13.7 (2) | 13.15 (2) | - |
| Sodium (median, IQR) | 136 (5) | 135 (4) | - |
| Potassium (median, IQR) | 3.8 (0.6) | 4 (0.5) | - |
| Creatinine (median, IQR) | 94.5 (38) | 85 (27.7) | - |
| Glucose (median, IQR) | 109 (32) | 114 (67) | - |
| C-reactive Protein (median, IQR) | 61 (124) | 91.8 (128) | - |
| **Drug Therapy** |  |  |  |
| Hydroxychloroquine (N, %) | 25 (96.2) | 11 (100) | 36 (97.3) |
| Lopinavir/Ritonavir (N, %) | 12 (46.2) | 5 (45.5) | 17 (45.9) |
| Tocilizumab (N, %) | 5 (19.2) | 5 (45.5) | 10 (27) |
| Antibiotics (N, %) | 9 (34.6) | 10 (90.9)* | 19 (51.4) |
| Corticosteroids (N, %) | 8 (30.8) | 10 (90.9)* | 18 (48.6) |
| **Outcomes** |  |  |  |
| Pneumonia (N, %) | 25 (96.2) | 11 (100) | 36 (97.3) |
| ICU (N, %) | - | 4 (36.4) | 4 (10.8) |
| Days from hospitalization to ICU (N, %) | - | 9.5 (5-12) | 9.5 (5-12) |
| Days in ICU (N, %) | - | 15 (15-22) | 15 (15-22) |
| Non-mechanical Ventilation (N, %) | - | 11 (100) | 11 (29.7) |
| Mechanical Ventilation (N, %) | - | 2 (18.2) | 2 (5.4) |
| Nosocomial Co-Infection (N, %) | - | 2 (18.2) | 2 (5.4) |
| Mortality (N, %) | - | 5 (45.5) | 5 (13.5) |
| Days of hospitalization (mean, range) | 8.2 (2-16) | 18.4 (3-47) | 11.2 (2-47) |

\*SOTR: solid organ transplant recipient; SpO2 < 94%: pulse oximetry below 94%; ICU: intensive care unit

Acute blood samples were collected in all patients upon hospital admission (day 0, recruitment). Subsequent blood draws were performed at day 3 and 7 post-recruitment in mild versus severe patients: (21 (80.8%) vs. 8 (72.7%) and 14 (53.8%) vs. 8 (72.7%), respectively. COVID-19 survivors were also followed during the convalescence phase, and an additional blood sample was collected from 22 (84.6%) mild vs. 6 (54.5%) severe patients, with a mean time of 46 days post-recruitment.

### Impaired T Cell Activation and Enhanced Inflammation in Severe COVID-19

Transcriptomic and proteomic profiling have proven powerful tools to stratify COVID-19 patients according to disease trajectory. Importantly, these approaches have been used to uncover key inflammatory and immunological pathways in mild versus severe COVID-19^3,4,11,17,42,43^. To capture both transcriptional programs and cytokine landscapes that distinguish mild from severe COVID-19 trajectories in the BACO cohort, we performed bulk RNA-sequencing on the whole peripheral blood from these patients followed by multiplex Olink proteomics of paired serum samples.

Differential gene expression analysis was used first to investigate the changes in gene expression between mild and severe COVID-19 patients. A heatmap of average gene expression differences in severe patients relative to mild cases showed that most transcriptional changes occurred during the acute phase, especially at days 3 and 7 post-recruitment (Figure 1A). Figure 1B shows Venn diagrams illustrating the overlap and distribution of upregulated and downregulated genes in severe versus mild COVID-19 patients at days 0, 3, and 7 post-recruitment. Severe patients showed an upregulation of 151 genes and downregulation of 364 genes compared to mild patients at baseline. By day 3, the number of differentially expressed genes (DEGs) in severe cases drastically increased, with 2639 upregulated and 2978 downregulated genes compared to mild cases. These differences started to progressively decrease over time, with 2100 upregulated and 2286 downregulated genes at day 7 post-recruitment, while transcriptomic differences were no longer significant by day 46, aligning with a shift toward disease resolution. Gene Ontology (GO) enrichment analysis further identified phenotype-specific differences in mild versus severe patients (Figure 1C). Data showed that mild COVID-19 patients had an early upregulation of antigen processing and presentation, as well as antigen assembly via MHC class II complex signatures. Consistently, the top enriched GO categories at day 3 and 7 post-recruitment in mild cases included T cell receptor complex, adaptative responses and ribosome biogenesis. In contrast, severe patients exhibited an increased transcriptional pattern associated with autophagosome assembly and organization, interleukine-1 receptor activity and secretory granules at day 3 and 7 post-recruitment. These findings were consistent with the top 20 genes found to be up- and down- regulated in severe cases over time: classical genes such as GPR84 and IL4R, associated with a pro-inflammatory state; or SRXN1, involved in oxidative stress, were upregulated while genes involved in T cell immunity such as CD27 and antigen presenting HLA molecules, such as HLA-DRA, HLA-DPA1 and HLA-DPB1 were downregulated at day 0 (Figure 1D and Supplementary Figure 1A). Interestingly, an early upregulation of LILRB3, a potent inhibitor of Fc receptor-mediated activation on myeloid cells was also found. This pro-inflammatory state was maintained over time, and most upregulated genes found at day 3 and 7 included classical genes such as IL1-R1, IL1-R2 and IL18-R1. In contrast, a diversity of T cell receptors including TRBV7-6, TRBV10-3, and TRAV27, were downregulated. Figure 1E and Supplementary Figure 1B show chord diagrams linking differentially expressed genes and their associated enriched GO categories in severe versus mild COVID-19 patients. Overall, our data suggested that upon SARS-CoV-2 infection, severe patients show an increased activation of pro-inflammatory and cellular stress-related transcriptional programs whereas mild cases exhibit a robust activation of immune responses.

**Figure 1.**
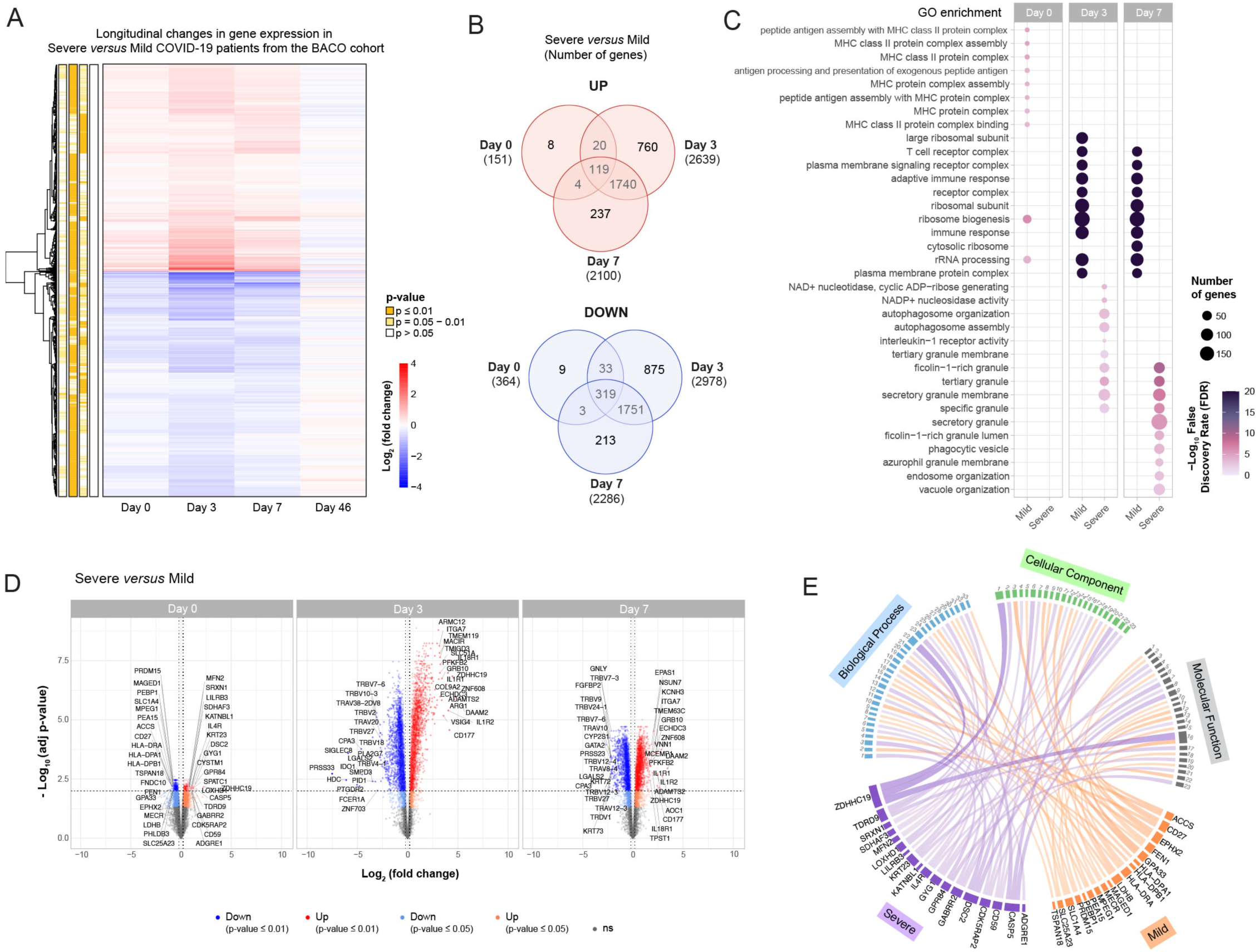
Longitudinal transcriptomic profile of mild and severe COVID-19 patients from the BACO cohort. **A)** Heatmap of differentially expressed genes (DEGs) in severe COVID-19 patients on days 0, 3, 7, and 46 post-recruitment. Average log_2_ fold changes in gene expression in severe versus mild patients are shown at each time point. Statistically significant differences are highlighted in yellow (empirical Bayes adjusted p-value ≤ 0.05, corrected for multiple testing using the Benjamini-Hochberg (BH) method). **B)** Venn diagram showing the number of DEGs upregulated (red) and downregulated (blue) in severe versus mild COVID-19 patients on days 0, 3, 7, and 46 post-recruitment. **C)** Gene ontology (GO) enrichment analysis of DEGs on days 0, 3 and 7 post-recruitment in mild and severe COVID-19 patients. Diagram shows the top 35 enriched GO terms across molecular function, cellular component and biological process categories. GO terms are ranked by their statistical significance, and dot size indicates overlap on number of genes. **D)** Volcano plots of upregulated (red) and downregulated (blue) genes in severe COVID-19 patients on days 0, 3, and 7 post-recruitment. Expression differences relative to mild patients and > 1.5-fold change are shown (log_2_ fold change ≤ 0.5, vertical dotted lines). Darker colors show genes with an adjusted p-value ≤ 0.01, while lighter colors show adjusted p-value ≤ 0.05. The names of the top 20 genes are shown. **E)** Chord diagram of Top 20 upregulated genes and their assigned GO pathways in mild and severe patients at day 0 post-enrollment. Specific pathways and corresponding GO accession numbers can be found in Supplementary Table 1.

To determine whether these transcriptional differences were reflected at the protein level, we next characterized the cytokine landscape of mild and severe patients using multiplex Olink proteomics. For this, a panel of 92 inflammation markers was used. Of these, 18 metabolites were under the limit of detection and excluded from downstream analysis. A heatmap of average cytokine changes in severe patients relative to mild cases is shown in Figure 2A. Consistent with the transcriptomic data, most proteomic changes occurred during the early acute phase, at day 0 and 3 post-recruitment, while few or no significant changes were observed at day 7, or the later convalescent timepoint, at day 46. Compared to mild patients, severe cases showed an early upregulation of pro-inflammatory cytokines, including OSM and CASP-8 as well as monocyte recruitment factors such as MCP-1 and MCP-3 (Figures 2B and 2C and Supplementary Figure 2). Differently expressed proteins drastically increased by day 3 post-recruitment with significant upregulation of inflammatory mediators such as IL7, OSM and IL18-R1. Interestingly, we also found increased levels of EN-RANGE, a well-known inflammatory mediator involved in myeloid cell activation, and previously linked to critical COVID-19 outcomes^44,45^. In parallel, proteins associated with T cell activation were downregulated, including CD8A, TNFRSF9, and IL12-β, consistent with the impaired lymphoid activation observed in the transcriptomic data. Additionally, levels of Fit3L, a cytokine associated with immune progenitor development, were markedly low in severe patients. Interestingly, canonical inflammatory cytokines (TNF, TNF-β) were also decreased in severe cases, perhaps reflecting regulatory mechanisms aimed at limiting excessive inflammation. By day 7 post-recruitment, the only significant differences found were consistent with a downregulation of markers associated with T cell impairment, including IL12-β, TNFRSF9 and TRANCE in severe patients.

**Figure 2.**
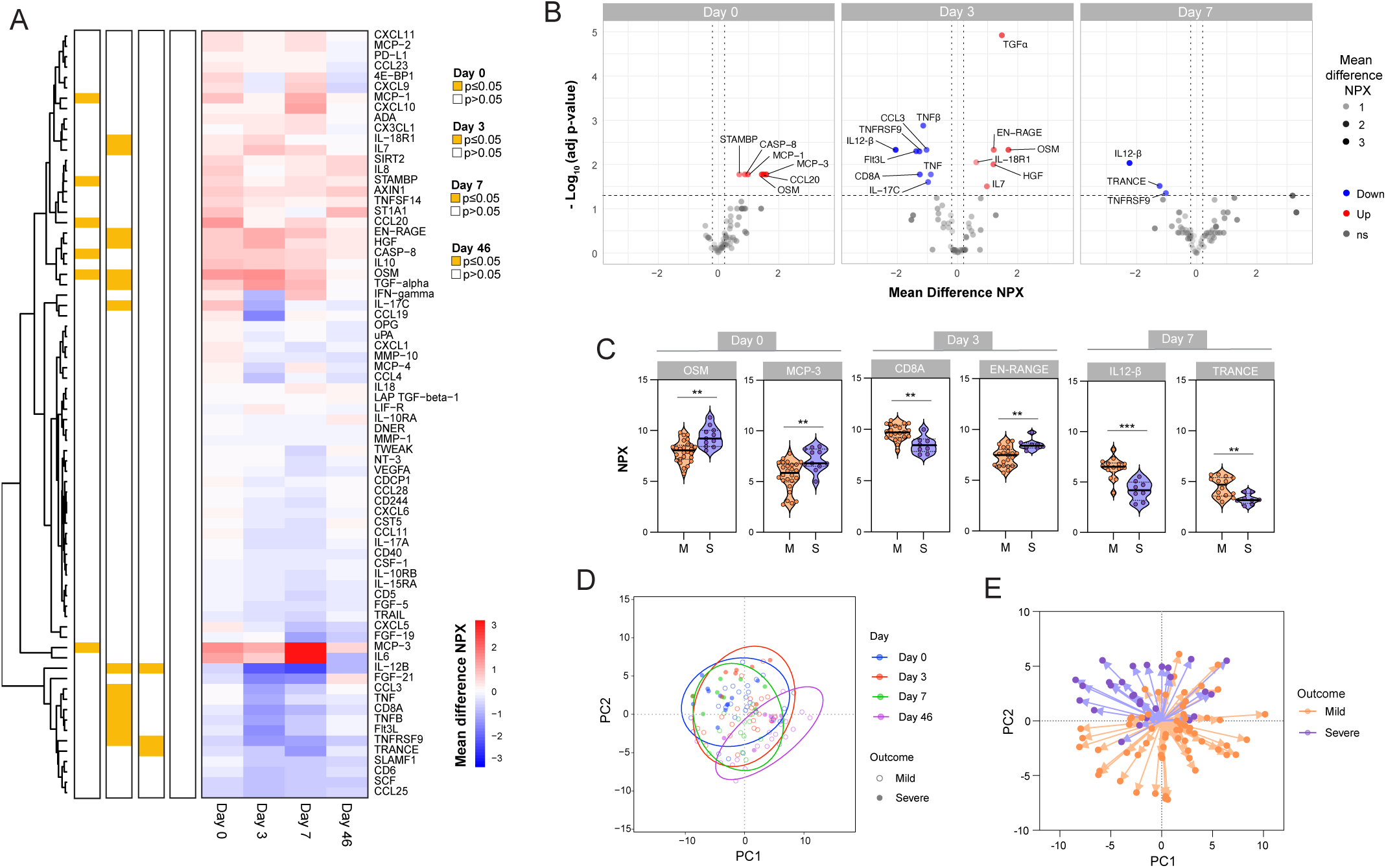
Differential cytokine signatures in mild and severe COVID-19 patients from the BACO cohort. **A)** Heatmap of differentially expressed cytokines in severe COVID-19 patients on days 0, 3, 7, and 46 post-recruitment. Olink-derived mean differences (NPX) in severe versus mild patients are shown at each time point. Statistically significant differences are highlighted in yellow (empirical Bayes adjusted p-value ≤ 0.05, corrected for multiple testing using the Benjamini-Hochberg (BH) method). **B)** Volcano plots of upregulated (red) and downregulated (blue) cytokines in severe COVID-19 patients on days 0, 3, and 7 post-recruitment. Protein expression differences relative to mild patients and NPX ≤ 0.5 (vertical dotted lines) are shown. **C)** Violin plots showing some differentially expressed cytokines in mild and severe COVID-19 patients on days 0, 3 and 7 post-recruitment. Mann-Whitney *U* test was performed to compare differences between mild vs. severe patients at each time point. Statistical significance was considered when p ≤ 0.05 (∗p < 0.05, ∗∗p < 0.01, ∗∗∗p < 0.001, ∗∗∗∗p < 0.0001). **D)** Principal Component Analysis (PCA) of Olink-based plasma cytokines according to day of recruitment (day 0: blue; day 3: red; day 7: green and day 46: purple). Empty circles represent mild cases, and filled circles represent severe cases. Ellipses show 95% confidence regions. **E)** PCA of Olink-based plasma cytokines according to COVID-19 severity (mild: orange; severe: purple) during the acute phase.

To identify longitudinal differences in cytokine regulation between mild and severe patients, we next performed Principal Component Analysis (PCA). PCA of cytokine profiles from the acute phase (days 0, 3, and 7) showed largely overlapping clustering, indicating minimal temporal separation and overall similar cytokine expression patterns early after infection. In contrast, convalescent samples collected on day 46 shifted distinctly away from acute-phase clusters, reflecting divergence as immune responses resolved (Figure 2D). Notably, when restricting the analysis to systemic cytokine levels during the acute phase, PCA revealed two opposing immunological trajectories for mild versus severe cases (Figure 2E). Together, our integrated transcriptomics and proteomics analysis confirmed two distinct immune phenotypes associated with COVID-19 disease severity, consistent with previously published studies^4,15,17^.

### Delayed Fc-Effector Activation and Early S2 Bias Characterize Severe COVID-19

Our RNA-seq analysis revealed early upregulation of LILRB3 in patients with severe COVID-19 (Figure 1D). Although not fully characterized, this gene has been implicated in regulating Fc receptor-mediated functions on myeloid cells^46–49^. These findings suggested that beyond T cells, Fc-driven mechanisms could also influence disease trajectory. Fc receptor activation occurs when antibodies engage viral glycoproteins and trigger innate effector functions through their Fc portion, providing protection that extends beyond neutralization^31,35,50^. Based on these observations, we next investigated two complementary questions in the BACO cohort: first, whether specific immune cell populations could shape clinical outcomes; and second, whether SARS-CoV-2 S-specific antibodies were capable of mediating Fc-effector functions and drive disease progression.

Our previously described computational cell type deconvolution approach was first applied^51^. For this, we leveraged transcriptional signatures from healthy PBMCs profiled by bimodal protein-RNA measurements with Cellular Indexing of Transcriptomes and Epitopes by Sequencing (CITE-Seq) at the single-cell level^51^. This method encompassed 37 cell types and subtypes spanning both lymphoid and myeloid compartments. A false discovery rate (FDR) < 0.05 (adjusted p-value,

Fisher’s exact test) was used to deconvolve the bulk RNA-seq data and identify cell type specific transcriptional signatures differentially enriched in the blood of the BACO cohort patients -mild versus severe- during the early timepoints (day 0, 3 and 7). Results revealed an early and significant enrichment of B and T cell lineages including plasmablasts, memory B cells and proliferating CD4+ and CD8+ T cells in mild patients, as compared to severe, at day 0 (Figure 3A). Interestingly, classical-CD14+ monocytes were also enriched in mild cases, whereas non-classical-CD16+ monocytes predominated in severe cases, reflecting distinct inflammatory dynamics early upon hospital admission. This initial response was followed by a broader expansion of immune cell populations at day 3 and 7 post-recruitment including both innate and adaptative immune cell populations in mild cases, whereas enriched cell populations in severe patients were largely restricted to monocyte and DCs. Polar plots illustrating the magnitude and temporal enrichment of these cellular changes are shown in Figure 3B. To further estimate cell type proportions across disease phenotypes, we performed a complementary deconvolution analysis using the UniCell Deconvolve framework ^52^. This independent approach produced results broadly consistent with our initial analysis, with only minor differences likely attributable to the distinct reference signatures used (Figure 3C and 3D and Supplementary Figure 3).

**Figure 3.**
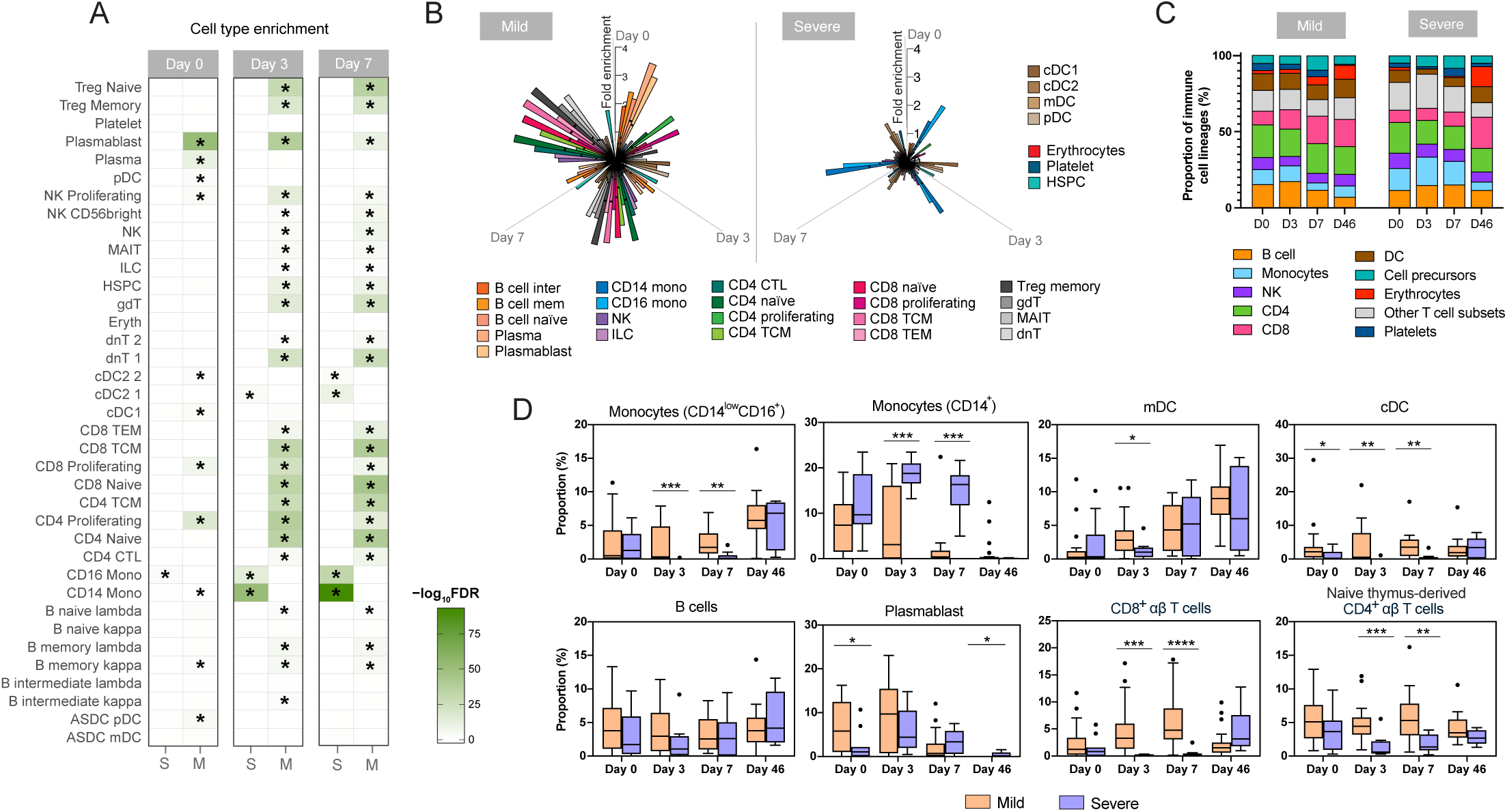
Cell type deconvolution reveals distinct immune populations associated with COVID-19 severity. **A)** Main cell types enriched in the peripheral blood of mild and severe COVID-19 patients on days 0, 3 and 7 post-recruitment. Asterisk indicates False Discovery Rate (FDR) < 0.05 (Fisher’s exact test, cell type reference from ref.^87^). **B)** Fold change enrichment of gene sets in mild and severe COVID-19 patients after cell type deconvolution on days 0, 3 and 7. Bar indicates fold change enrichment for each cell type at each time point over day 0. **C)** Bar plot of the proportion of immune cell types in mild and severe COVID-19 patients after cell type deconvolution by the UniCell Deconvolve (UCD) method. **D)** Box-and-whisker diagrams showing the relative abundance of immune cell types by the UCD method in mild and severe COVID-19 patients. Box indicates interquartile range (IQR, Q1-Q3), with horizontal line showing the median and vertical lines indicating minimum and maximum. Mann-Whitney *U* test was performed to compare differences between mild vs. severe patients at each time point. Statistical significance was considered when p ≤ 0.05 (∗p < 0.05, ∗∗p < 0.01, ∗∗∗p < 0.001, ∗∗∗∗p < 0.0001).

We next investigated antibody specificity and Fc-mediated activity, including antibody-dependent cellular cytotoxicity (ADCC) and antibody-dependent cellular phagocytosis (ADCP) of the patient’s polyclonal sera. Because antibodies directed towards the S1/RBD versus the S2 subunit have been previously shown to exert different functional properties^26,53–55^, we first compared antibody induction between the S1/RBD- and S2- subdomains according to disease phenotype (Figure 4A and Supplementary Figure 4A). For this, we longitudinally profiled the IgG binding antibody response against SARS-CoV-2 S1, RBD and S2 domains in mild and severe patients using ELISA assay. Results showed an increase in S1- and RBD- specific antibody titers in patients with mild disease early after infection, especially at days 0 and 3 post-recruitment. In contrast, severe patients exhibited elevated levels of antibodies targeting the more conserved S2 domain. Consistently, S2/S1 and S2/RBD ratio analysis showed proportionally higher anti-S2 responses in severe patients, indicating an early skewing toward conserved epitopes (Figure 4B). To exclude a potential contribution of pre-existing immunity from seasonal HCoVs, we also measured systemic antibody responses against full-length S and S1/S2 subdomains of HCoV-OC43 (beta-CoV) and full-length S of HCoV-229E (alpha-CoV). No significant differences were found between mild versus severe cases (Supplementary Figure 4B). Next, we quantified ADCC and ADCP activity using our in-house modified bioreporter assay based. For this, an A549 cell line stably expressing SARS-CoV-2 full-length S protein was generated, as previously described^34,56,57^ (Supplementary Figure 4C and 4D). As shown in Figure 4C, both groups showed comparable levels of antibodies with ADCC activity throughout the entire follow-up period. In contrast, patients with mild disease showed higher levels of ADCP activity at day 3 as compared to severe cases, whereas severe patients exhibited delayed induction, reaching levels similar to mild cases only by days 7 and 46. These results suggest that similar to binding antibody responses, severe COVID-19 patients displayed delayed kinetics of induction rather than impaired Fc-mediated antibody responses.

**Figure 4.**
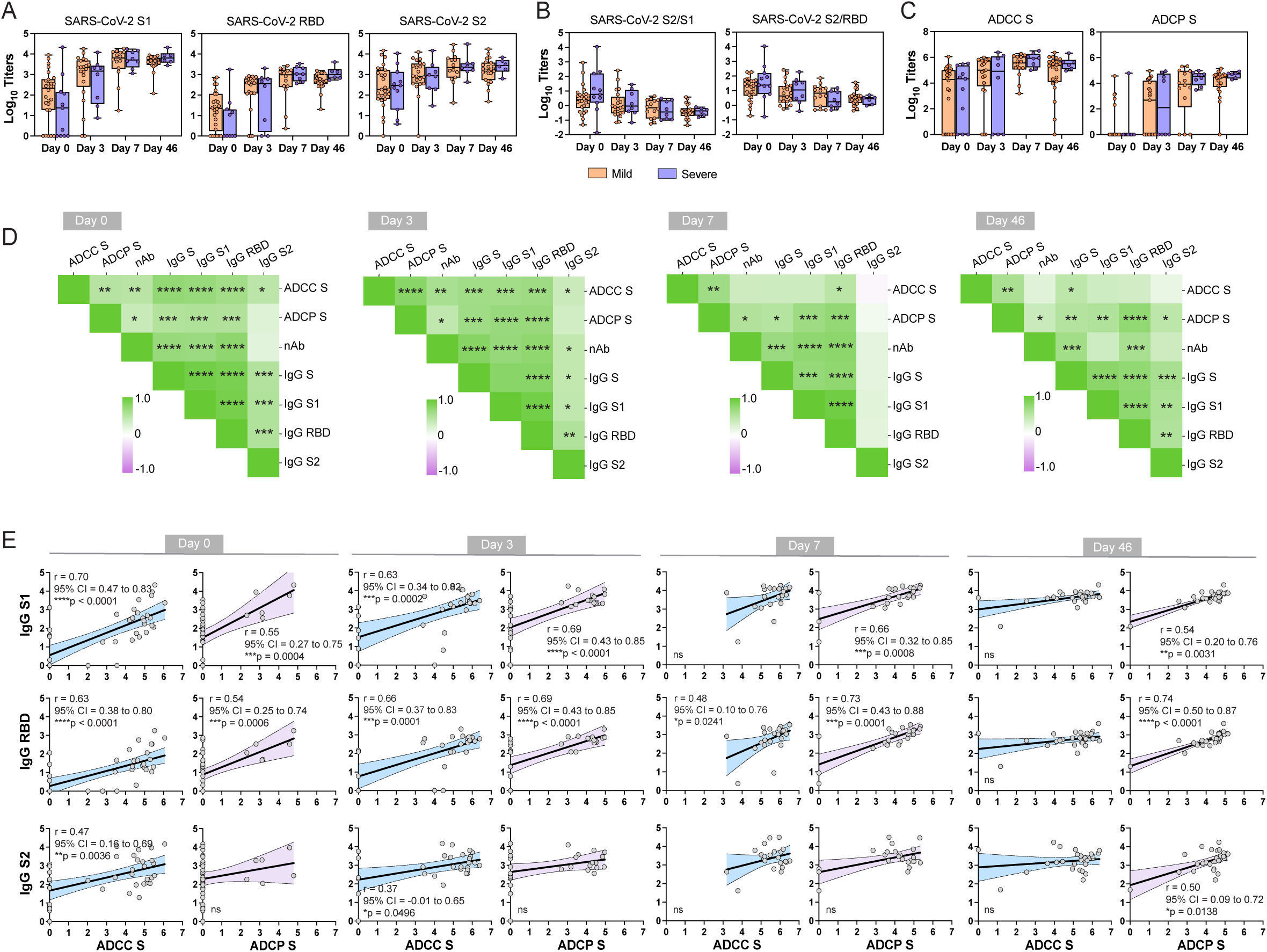
Mild COVID-19 patients show an early increase in ADCP responses associated with anti-S1 antibodies. Box-and-whisker plots showing **(A)** the binding antibodies titers against SARS-CoV-2 S1, RBD and S2 spike antigens **(B)** the ratios between S1/S2 and RBD/S2 binding titers and **(C)** the Fc-receptor antibody-mediated ADCC and ADCP activities against SARS-CoV-2 full-length spike in mild and severe COVID-19 patients. Box indicates interquartile range (IQR, Q1-Q3), with horizontal lines showing the median and vertical lines indicating minimum and maximum. Mann-Whitney *U* test was performed to compare differences between mild vs. severe patients at each time point. Statistical significance was considered when p ≤ 0.05. **D)** Heatmap of Spearman correlation matrices between SARS-CoV-2 antibody titers (ADCC, ADCP, nAb and IgG binding titers against S, S1, S2 and RBD) at days 0, 3, 7 and 46 post-recruitment. Statistically significant correlations in the underlined intersections are indicated with asterisk (∗p < 0.05, ∗∗p < 0.01, ∗∗∗p < 0.001, ∗∗∗∗p < 0.0001). **E)** Spearman correlations between binding IgG antibodies against S1, RBD or S2 domains and functional ADCC (blue) or ADCP (purple) antibodies at each timepoint. Each dot represents a sample. Shown are the Spearman r coefficient, 95% confidence interval (CI) and p value (∗p < 0.05, ∗∗p < 0.01, ∗∗∗p < 0.001, ∗∗∗∗p < 0.0001, ns, not significant). Data is also shown in Supplementary Table 2.

Given the parallel dynamics observed for S1/S2 antibody specificity and ADCP activity across disease outcomes over time, we next performed correlation analyses to investigate the relationship between antibody specificity and functional activity. Figure 4D shows Spearman correlation matrices illustrating the relationships between ADCC, ADCP and neutralizing activity and IgG antibody binding to different SARS-CoV-2 antigens. As expected, antibodies with neutralizing activity showed strong correlation with anti-S1 and RBD domains while poor correlation was observed with anti-S2 antibodies. Moreover, neutralizing antibodies also showed a significant correlation with both ADCC and ADCP activities. Consistently, S1- and RBD-specific antibodies also showed significant associations with ADCC and ADCP at days 0 and 3 post-recruitment, underscoring their central role in driving both neutralizing and Fc-effector functions. In contrast, anti-S2 antibodies showed only poor correlation with ADCC activity while no correlation was found with ADCP activity at day 0 and 3 post-recruitment. Individual Spearman correlation coefficients and corresponding p-values for Fc functional activity and antibody features at each time point are shown in Figure 4E and Supplementary Table 3. Overall, these findings suggest that antibody functionality including Fc-effector functions might be primarily driven by responses targeting the S1 domain of the SARS-CoV-2 S protein in the BACO cohort. The earlier induction of S1-driven Fc responses in mild cases, contrasted with a delayed onset in severe cases, highlighting a potential protective role of a timely antibody-mediated Fc activity. Conversely, severe patients showed earlier induction of S2 antibodies, but these contributed little to Fc-effector functionality, further underscoring the importance of rapid S1-directed responses for favorable outcomes.

### Early enhanced Fc-effector monocyte phagocytosis by S1-specific antibodies is linked to mild COVID-19 disease

To further dissect the role of anti- S1 and S2 antibodies in mediating Fc-effector functions, we next performed an integrative system serology approach. Using an Fc-binding protein array, we quantified the ability of antibodies targeting S1 versus S2 to engage in activating and inhibitory Fcγ receptors (FcγR2A, FcγR2B, FcγR3A, and FcγR3B). This approach serves as a valuable proxy to assess domain-specific activity to drive Fc-mediated effector functions. The two common activating allelic variants of FcγR2A (H/R) involved in ADCP activity, and FcγR3A (F/V) involved in ADCC function, were tested. A heatmap of the binding data according to disease phenotype is shown in Figure 5A. Overall, mild patients had a stronger Fcγ-receptor binding profile of SARS-CoV-2 antibodies as compared to severe cases (Figure 5A). Interestingly, a delay on S binding responses was observed in severe cases: mild patients exhibited stronger Fc-binding early (days 0 and 3) especially for anti-S1 antibodies, whereas severe patients progressively increased their Fc-receptor engagement, reaching levels comparable to mild cases by days 7 and 46 post-recruitment. (Figure 5A-C).

**Figure 5.**
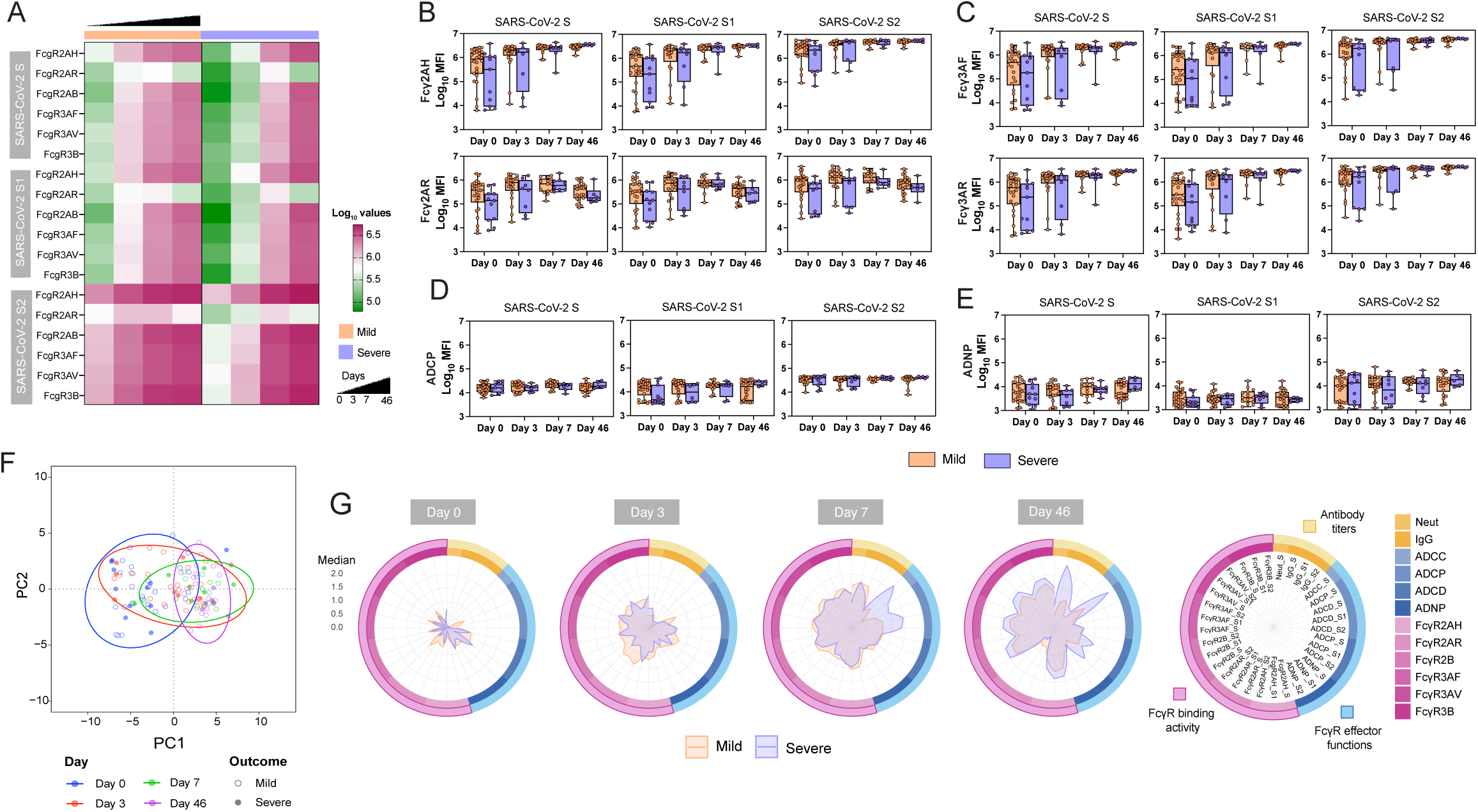
Severe COVID-19 patients exhibit a delay in Fc-mediated phagocytosis functions. **A)** Heatmap of FcγR-binding antibody responses to SARS-CoV-2 S antigens in mild and severe COVID-19 patients. Binding activity of antibodies to FcγRs was measured for SARS-CoV-2 full-length S, S1 and S2 antigens at days 0, 3, 7, and 46 post-recruitment. Box-and-whisker diagrams showing **(B)** FcγR2AH and FcγR2AR-binding antibody responses **(C)** FcγR3AV and FcγR3AF-binding titers, **(D)** functional monocyte antibody-dependent cellular phagocytosis (ADCP) responses and **(E)** functional antibody-dependent neutrophil phagocytosis (ADNP) responses against SARS-CoV-2 full-length S, S1 and S2 antigens in mild and severe COVID-19 patients. Box indicates interquartile range (IQR, Q1-Q3), with horizontal line showing the median and vertical lines indicating minimum and maximum. Mann-Whitney *U* test was performed to compare differences between mild vs. severe patients at each time point. Statistical significance was considered when p ≤ 0.05. **F)** PCA of antibody responses according to day of recruitment (day 0: blue; day 3: red; day 7: green and day 46: purple). Empty circles represent mild cases, and filled circles represent severe cases. Ellipses show 95% confidence regions. **G)** Spider plot of the antibody landscape (IgG antibody titers, FcγR-binding activity and FcγR-effector functions) in mild and severe COVID-19 patients. Median percentile for each antibody feature is shown at each timepoint and for each SARS-CoV-2 S antigen.

Given that ADCP activity can be mediated by different myeloid immune cells, we next assessed the functional capacity of SARS-CoV-2-specific antibodies to mediate antibody-dependent phagocytosis by human monocytes (ADCP) or primary neutrophils (ADNP). Results showed that anti-S1 antibodies with ADCP activity tend to be higher in mild patients at day 0 and 3 post-recruitment while no differences were observed for anti-S2 antibodies (Figure 5D). Similarly, antigen-specific ADNP activity did not strongly differ between mild and severe patients (Figure 5E). While only indicative, these data suggested a more prominent role for monocyte-mediated phagocytosis by S1-speficic antibodies, rather than neutrophil-driven responses, in early protection associated with mild COVID-19 disease. Overall, Fc-binding patterns and temporal dynamics were then explored using PCA of the full Fc-engagement dataset. (Figure 5F). PCA revealed partial separation between Fc-binding profiles at days 0, 3, 7, and 46. However, greater overlap was observed between profiles at days 0 and 3, as well as between days 7 and 46. Finally, and to see disease projection over time based on all the antibodies features measured, we represent overlapping spider plots according to disease phenotypes by centering our data to the median values for each antibody feature (Figure 5G). With this analysis, we aimed to define disease trajectory based on the global serological immune landscape of mild and severe COVID-19 patients. In general, mild patients showed early signatures of ADCP activity and strong binding to FcγR2 receptors as compared to severe cases. This was followed by a broad increase in FcγR-mediated functionality and binding activity in mild patients by day 3 post-recruitment, whereas severe patients exhibited delayed kinetics in the induction of these antibody features. By day 7, the immune profiles of both groups began to converge; however, severe cases showed a marked expansion of ADCC activity and complement deposition (Supplementary Figure 5A). During the convalescent phase, severe patients continued to display a broader and more robust range of antibody functions compared to mild cases. Taken together, our results suggest that severe COVID-19 is associated with a delayed development of binding, neutralizing, and Fc-effector antibody responses. Importantly, our study also describes the role of S1-specific antibodies in mediating Fc-effector functions and underscores the importance of an early induction of these S1-specific Fc-active antibodies to promote favorable disease outcomes.

## DISCUSSION

Outcomes of SARS-CoV-2 infection range from complete protection to severe, life-threatening disease^4,17,39^. Neutralizing antibodies contribute to protection^58,59^ but emerging evidence shows that Fc-mediated antibody effector functions, mucosal humoral immunity, and innate myeloid regulation are central to early antiviral defense^60–68^. Yet, how these pathways integrate in the earliest stages of infection to determine disease severity remains unclear. In this study, we used samples from SARS-CoV-2 immunologically naïve patients –The BACO cohort– to identify early immune signatures of COVID-19 outcomes. By integrating multi-omics approaches and system serology analyses, we found two distinct disease trajectories corresponding to mild and severe outcomes. Mild patients exhibited higher levels of Fc-effector phagocytosis early upon infection as compared to patients with severe disease. Interestingly, these differences in Fc-effector functionality were primarily driven by S1-specific antibodies rather than antibodies against the more conserved S2 domain. Overall, these findings highlight the importance of the early induction of S1-specific, Fc-active antibodies in promoting favorable disease outcomes and underscore the importance of eliciting functionally potent humoral responses in future therapeutic strategies.

An important hallmark of severe COVID-19 is the profound dysregulation of innate and myeloid cell states characterized by inflammatory monocyte expansion, impaired antigen presentation, and myeloid immunosuppression^69–71^. Our transcriptomic analysis recapitulated this pattern revealing profound dysregulation of multiple immune pathways in severe versus mild patients: impaired T cell activation, decreased antigen processing and presentation; and enhanced inflammation, protein lipidation and secretory granule production (Figure 1C). This was accompanied by altered expression of several canonical genes associated with these transcriptional programs, including multiple TCR chains, HLA molecules, ZDHHC19 and CD177^17,72–76^ (Figure 1D and E). Consistently, the analysis of plasma cytokines showed enhanced levels of several proinflammatory markers in severe COVID-19 including monocyte chemoattractant factors such as MCP-1 and MCP-3, OSM, IL7, IL18-R1 and EN-RANGE (Figure 2B and C). This elevated plasma levels of MCP-1 and MCP-3 aligned with the selective enrichment of monocyte populations, particularly classical CD14⁺ monocytes, as revealed by our two independent cell-type deconvolution approaches. Classical CD14⁺ monocytes are typically pro-inflammatory and have higher phagocytic activity as compared to non-classical monocytes. However, despite their increased abundance in severe COVID-19, BACO patients exhibited reduced Fc-effector phagocytosis, suggesting that classical monocytes, while expanded, may be functionally impaired. This could be potentially due to inhibitory signals. It is particularly interesting that we found an early upregulation of the LILRB3 gene in severe patients (Figure 1D and Supplementary Figure 1C). This receptor (also known as ILT5 or CD85a) has been previously described as an immunoregulatory checkpoint expressed on myeloid cells including monocytes and neutrophils^46–49,77–79^. While the specific role is not clear, inhibitory receptors of the LILRB family have previously implicated on the immune response to pathogens^78,80^.This has been documented in HIV-1, where LILRB1/2 engagement promotes immune evasion^81^, and in Zika virus, where LILRB polymorphisms influence infection susceptibility^82^. Although direct evidence in SARS-CoV-2 remains emerging, we speculate that LILRB-associated inhibitory pathways may contribute to Fc-immunity failure and severe COVID-19 by preventing an efficient clearance of antibody-opsonized virus and sustained hyperinflammation. Further studies will be needed to establish a causative link between LILRB3 expression and COVID-19 disease progression.

Our findings are highly relevant since Fc-effector functions including ADCC and ADCP activities have been shown to contribute to protection against several viral infections^35–37,83,84^. For influenza virus, most Fc-active antibodies recognize the highly conserved stem region of the hemagglutinin protein. However, for SARS-CoV-2, the specificity/functionality axis is less clear. Although Fc-effector antibodies targeting the S2 domain have been previously described^31,85,86^, our data indicated that antibodies targeting the S1 subunit (and therefore likely neutralizing) can similarly trigger Fc-mediated antibody functions. Notably, mild patients from the BACO cohort displayed higher levels of anti-S1 antibodies early after infection, whereas severe patients mounted higher anti-S2 responses. Importantly, these early anti-S2 antibodies were poor inducers of ADCP activity, suggesting that an early S2-skewed response may be functionally suboptimal. The functional interplay between S1/S2 was further supported by our FcR multiplex arrays, which showed preferential binding of S1-specific antibodies to FcR mediating phagocytosis in mild versus severe patients of the BACO cohort. Therefore, beyond neutralization, S1-specific antibodies can potentially exert complementary antiviral immunity by recruiting innate immune mechanisms. These discoveries inevitably raise important questions about immune imprinting, not only in shaping epitope specificity but also in determining antibody functionality. Severe disease was characterized by an early dominance of S2-biased, low-effector antibodies, a pattern consistent with immune memory recall of conserved epitopes. Such imprinting toward conserved S2 regions may limit the induction of functionally potent S1-specific antibodies and potentially undermine Fc-effector activity during early infection. The role of anti-S1 antibodies in protection is further supported by complementary observations from the CIDS cohort, a sister study to evaluate determinants of SARS-CoV-2 virus transmission that we recently published^68^. In this study, we showed that protection from infection upon SARS-CoV-2 exposure was characterized by pre-existing, S1-focused, and likely Fc-competent, antibody responses.

In summary, our work demonstrates that early functional humoral immunity, particularly the rapid induction of S1-specific Fc-active antibodies, is a key determinant of COVID-19 outcomes. Mild disease was characterized by timely S1-driven phagocytic activity and coordinated innate responses, whereas severe disease exhibited early S2-biased, low-effector antibodies, innate dysregulation, and inhibitory LILRB3-associated myeloid states. Altogether, our work provides foundational knowledge and a conceptual framework for further investigations into Fc-effector–associated responses and other immunological mechanisms in COVID-19. Future studies with larger and more diverse cohorts will be necessary to validate and extend these observations.

Limitations: we acknowledge several limitations. First, the BACO cohort consists of 37 hospitalized COVID-19 patients from Spain. Although the BACO cohort captures early immune responses in immunologically naïve individuals, the sample size limited our ability to fully stratify patients by additional factors such as age or comorbidities, all of which can influence disease outcomes. Second, we provide evidence supporting the potential role for LILRB3 in shaping myeloid dysfunction during severe COVID-19. However, our observations are correlative, and mechanistic studies are needed to determine causality.

## MATERIALS AND METHODS

### The BACO Cohort

The BACO cohort is a prospective human cohort study of COVID-19 disease carried out during the first pandemic wave (March–May 2020) of SARS-CoV-2 in Barcelona (Spain)^39,40^. A positive COVID-19 case was defined according to international guidelines when a nasopharyngeal (NP) swab tested positive for SARS-CoV-2 by reverse transcriptase quantitative polymerase chain reaction (RT-qPCR) upon hospital admission. All patients or their legally authorized representatives provided informed consent prior to sample and data collection. Serum samples from patients were collected at the enrollment in the study (baseline), and at days 3 and 7 post-enrollment. A convalescence sample was collected from survivors after recovery and hospital discharge with a mean time of 46 days (range, 30-56 days). A total of 37 serum samples were collected on day 0, followed by 29 samples on day 3, 22 samples on day 7, and 28 samples during the convalescence phase. All samples were stored at -80°C. Data on demographics, including age and sex, comorbidities, clinical signs and symptoms, interventions, and outcomes have been previously described in Aydillo *et al.,* 2021^39^ and Escalera *et al*., 2024^40^ and can also be found in Table 1. Severity of COVID-19 was assigned following a described severity scale based on oxygen saturation (SpO2), presence of pneumonia/imaging, oxygen support defined as use of high-flow nasal cannula (HFNC), non-rebreather mask (NRB), bilevel positive airway pressure (BIPAP) or mechanical ventilation (MV); and kidney (creatinine clearance, CrCl) and liver (alanine aminotransferase, ALT) function^41^: mild (SpO2 > 94% AND no pneumonia), moderate (SpO2 < 94% AND/OR pneumonia), severe (use of HFNC, NRB, BIPAP or MV AND no vasopressor use AND CrCl >30 AND ALT < 5x upper limit of normal) and severe with end-of organ disease (Use of HFNC, NRB, BIPAP or MV AND vasopressor use OR CrCl >30 or new HD OR ALT < 5x upper limit of normal). The study protocol was approved by the Institutional Review Board of University Hospital of Bellvitge, Barcelona, Spain, and by the Icahn School of Medicine at Mount Sinai, New York, US.

### Cell lines

VeroE6 cells, human embryonic kidney 293T (HEK293T) cells and human lung epithelial (A549) cells were originally purchased from the American Type Culture Collection (ATCC). A master cell bank was created for each cell line and early-passage cells were thawed in every experimental step. Cells were maintained in complete Dulbecco’s modified Eagle’s medium (cDMEM) with glucose, L-glutamine, and sodium pyruvate (Corning, 10-017-CV) supplemented with 10% fetal bovine serum (FBS, GIBCO,16140071), non-essential amino acids (Corning, 25-025-CI), penicillin (100 UI/mL), streptomycin (100 UI/mL) (Corning, 30-002-CI) and normocin (100 ug/mL) (InvivoGen, ant-nr-1) to prevent mycoplasma infection. Cells were grown at 37°C in 5% CO_2_.

### Virus strains

SARS-CoV-2 USA-WA1/2020 isolate was initially obtained from BEI Resources (Cat. #NR-52281). Viral stock was produced by infecting VeroE6 cells at a MOI of 0.01. Infected cells were maintained in infection media (DMEM with glucose, L-glutamine, and sodium pyruvate supplemented with 2% FBS, non-essential amino acids, HEPES, penicillin (100 UI/mL) and streptomycin (100 UI/mL)) at 37°C in 5% CO_2_. Infected cells were monitored by microscopy and cell-infected supernatants were collected at day 2 post-infection when cytopathic effect was observed. Viral supernatants were clarified of cell debris by spin and aliquots were stored at - 80°C. Viral stock was sequence-confirmed before performing any experiments.

### Recombinant proteins

All recombinant proteins used in this study were purchased from Sino Biological: SARS-CoV-2 full-length spike (S) protein (Cat. #40589-V08H4), seasonal HCoV-OC43 full-length S (Cat. #40607-V08B), seasonal HCoV-229E full-length S (Cat. #40605-V08B), SARS-CoV-2 S1 domain (Cat. #40591-V08H), SARS-CoV-2 S2 domain (Cat. #40590-V08H1), SARS-CoV-2 RBD subdomain (Cat. #40592-V08H80), HCoV-OC43 S1 domain (Cat. #40607-V08H1) and HCoV-OC43 S2 domain (Cat. #40607-V08B1). All recombinant proteins were stored at -80°C until use.

### Enzyme-linked immunosorbent assay (ELISA)

ELISA assays were performed as previously described^39,40^. Briefly, Immulon 4 HBX 96-well micro-titer plates (Thermo Fisher Scientific) were coated overnight at 4°C with 50 µl of the corresponding recombinant protein at a concentration of 2 µg/mL. The next day, plates were washed three times with phosphate-buffered saline (PBS; Corning) containing 0.1% Tween-20 (PBS-T, Fisher Scientific) using an automatic plate washer (BioTek). After washing, the plates were blocked for 1 h at room temperature (RT) with 200 µl/well of 3% (w/v) non-fat milk powder diluted in PBS-T. The blocking solution was removed, and 100 µl of serum samples diluted (starting concentration of 1:80 and serially diluted three-fold) in PBS-T containing 1% (w/v) non-fat milk was added to the wells and incubated for 1 h at RT. The plates were washed three times with PBS-T and 100 µl of anti-human IgG-HRP antibody (Fc-specific) (Sigma, Cat. A0170) at a dilution of 1:20,000. All secondary antibodies were diluted in PBS-T containing 1% (w/v) non-fat milk and incubated for 1h at RT. The plates were washed four times with PBS-T with shaking using the plate washer and 100 mL of TMBE (Rockland) was added to all wells for 10 min. The reaction was stopped by the addition of 100 mL of sulfuric acid per well. Optical density (OD) at a wavelength of 450 nm was read using Synergy 4 (BioTek) plate reader. OD values of samples were adjusted by subtracting the average of the blank plus three times the standard deviation of the blank. Area under the curve (AUC) values were computed by plotting normalized OD values against the reciprocal serum sample dilutions in GraphPad Prism. The assay was done one per sample due to the limited amount of sample.

### Microneutralizations

Microneutralization assays for antibody characterization were performed as previously described^39,89^. Briefly, VeroE6 cells were seeded in a 96-well plate and cultured in cDMEM. The following day, heat-inactivated serum samples (dilution of 1:10) were serially diluted three-fold in 1x minimum essential medium (MEM) with 2% FBS in a final volume of 200 µl. Then, 80 µl of serum dilution and 80 µl of SARS-CoV-2 virus diluted to 600 Tissue Culture Infectious Dose 50% (TCID_50_) were added to a new 96-well plate and incubated for 1 hr at 37°C. Next, cDMEM media was removed from cells and 120 µl of virus-serum mixture was added to the cells. The cells were incubated at 37 °C for 1 hr. After 1 hr incubation, virus-serum mixture was removed from the cells and 100 µl of the corresponding serum dilutions and 100 µl of 1x MEM with 2% FBS were added to the cells. Then, cells were incubated for 24 hr and fixed with 10% paraformaldehyde (Polysciences) for 24 hr at 4°C. Following fixation, cells were washed with phosphate-buffered saline (Corning) with Tween-20 (Fisher) (PBST) and permeabilized with 0.1% Triton X-100 (Fisher) for 15 min at room temperature (RT). The cells were washed three times with PBST and blocked with 3% milk in PBST for 1 hr at RT. Then, cells were incubated with anti-SARS nucleocapsid antibody (1C7C7, kindly provided by Dr. Moran at ISMMS) at a dilution of 1:1,000 in 1% milk in PBST and incubated for 1 hr at RT. The cells were washed three times with PBST. Then, cells were incubated with goat anti-mouse immunoglobulin G-horseradish peroxidase (IgG-HRP) (Abcam, Cat. ab6823) at a dilution of 1:10,000 in 1% milk in PBST and incubated for 1 hr at RT. The cells were washed three times with PBST and TMB ELISA peroxidase substrate (Rockland) was added. After 15 min incubation, sulfuric acid 4.0 N (Fisher) was added to stop the reaction, and the readout was done using a Synergy H1 plate reader (BioTek) at an OD450. Neutralizing antibodies titers were plotted as reciprocal dilution of serum and percentage of inhibition of virus using GraphPad Prism 9. Nonlinear regression curve fit analysis over the dilution curve was performed to calculate the half-maximum inhibitory concentration (IC_50_) values.

### Generation of A549 cell line stably expressing HexaPro-Spike

The open reading frame (ORF) encoding the HexaPro-stabilized S protein of the Wuhan SARS-CoV-2 strain^90^ was cloned into pLVX-IRES-Puro lentivirus vector (Takara). HEK293T cells were co-transfected with pLVX-IRES-Puro-HexaPro-S, lenti-gag/pol, and pMD2VSV-G using TransIT-LT1 (Mirus) according to the manufacturer’s guidelines. Supernatant was collected at 48 hrs post transfection and fresh media was added and collected at 60 hrs post transfection. Both supernatants were combined and spun at 1200 rpm for 10 min in a tabletop centrifuge (Eppendorf). The supernatant was filtered through a 0.45 µm cellulose acetate filter and aliquoted and frozen at -80 °C. A549 cells were seeded in a 6 well plate and fresh media, lentivirus and polybrene (1 µl of 12 mg/ml stock; Millipore, TR-1003G) were added. Four days post-transduction cells were selected with puromycin. Clonal population was generated, and clones were tested by indirect immunofluorescence using anti-spike antibody (KL-S-3A7)^88,91^.

### Antibody-dependent Cellular Cytotoxicity (ADCC) and Antibody-dependent Cellular Phagocytosis (ADCP) Bioassays

ADCC and ADCP reporter assays were performed using ADCC and ADCP Reporter Bioassay kits (Promega) with modifications^34^. A549-HexaPro S cells were seeded in a sterile white flat bottom tissue culture-treated 96-well plates at a density of 3.5 x 10^4^ cells/well. The following day, cells were washed with Opti-MEM 1X Reduced Serum Medium once and 25 µl of RPMI 1640 was added to each well. In a separate 96-well plate, serum samples were serially diluted 2-fold in Roswell Park Memorial Institute (RPMI) 1640 medium with a starting concentration of 1:25 and 25 µl of diluted serum was added to the cells. ADCC or ADCP effector cells (Jurkat T cells stably expressing nuclear factor of activated T cells (NFAT)-luciferase reporter and human FcγRIIIa or FcγRIIa, respectively) were diluted in RPMI 1640 medium and added to the wells (25 µl l/well) at a 1:1 final ratio (A549 : ADCC/ADCP ratio). Irrelevant serum was used as a negative control. After 5.5 hours of incubation at 37°C, 75 µl/well of Bio-Glo luciferase assay reagent was used to lyse the cell mix, and luminescence was measured using a Synergy H1 hybrid multimode microplate reader. The average background plus two standard deviations was used to discriminate between positive and negative values. Data were plotted in GraphPad Prism 9 software to calculate AUC.

### RNA isolation, library preparation, and sequencing

PAXgene blood samples were processed for total RNA extraction using the Agencourt RNAdvance Blood Kit (Beckman Coulter) on a BioMek FXP Laboratory Automation Workstation (Beckman Coulter) according to the manufacturer’s instructions. Concentration and RNA integrity number (RIN) of isolated RNA were determined using Quant-iT™ RiboGreen™ RNA Assay Kit (Thermo Fisher) and an RNA Standard Sensitivity Kit (DNF-471, Agilent Technologies, Santa Clara, CA, USA) on a Fragment Analyzer Automated CE system (Agilent Technologies), respectively. Subsequently, RNA-seq libraries were constructed from 300 ng of total RNA using the Universal Plus mRNA-Seq kit (Tecan Genomics, San Carlos, CA, United States) in a Biomek i7 Automated Workstation (Beckman Coulter). The transcripts for ribosomal RNA (rRNA) and globin were further depleted using the AnyDeplete kit (Tecan Genomics) prior to the amplification of libraries. Library concentration was assessed fluorometrically using the Qubit dsDNA HS Kit (Thermo Fisher), and quality was assessed with the Genomic DNA 50Kb Analysis Kit (DNF-467, Agilent Technologies). Preliminary sequencing of the libraries was performed using a MiSeq system (Illumina) to confirm library quality. Deep sequencing was subsequently performed using an S2 flow cell in a NovaSeq sequencing system (Illumina) (average read depth ∼30 million pairs of 2 × 95 bp reads) at the New York Genome Center.

### Bulk RNA-sequencing analysis

Illumina’s Real-Time Analysis (RTA) software was used for base-calling and quality scoring of RNA-sequencing (RNA-seq) data. Sequencing reads were processed and mapped to the human hg38 reference genome (Release 41 GRCh38.p13) with custom analysis scripts that combine publicly available tools as described before^92^. A combined matrix of mapped paired end read raw counts (genes x samples) was then obtained with featureCounts^93^ and used as input for differential gene expression (DGE) analysis in R v3.6.2. Prior to DGE analysis, gene counts were normalized to fragments per kb per million reads (FPKM) with RSEM with default settings for strand-specific data^94^. Genes with <1 FPKM in at least 50% of samples were removed from the analysis. Next, normalization factors were estimated using the trimmed mean of M-values (TMM) method, followed by voom mean-variance transformation^95^ to account for differences in coverage across samples. The data were inspected for potential confounders including the variables sex, site where sample was collected, RNA extraction batch and input RNA concentration, with Limma linear modeling^96^. Patient ID was used as a blocking variable to account for correlation between repeated measurements. Comparisons of Severe/SevereEOD against Mild/Moderate group were performed to determine changes in gene expression for each timepoint. For the longitudinal comparisons, each timepoint of sample collection within a group was compared to that group’s baseline reference (day 0). To determine genes with significant expression differences, Limma’s empiric Bayes adjusted p values were corrected for multiple testing using the Benjamini-Hochberg (BH) method (p ≤ 0.05).

### Gene ontology enrichment analysis

Gene ontology (GO) biological process (BP), molecular function (MF), and/or cellular component (CC) enrichment analyses of differentially expressed genes were performed using the gprofiler2 R v0.2.3 package^97^. The background gene set was restricted to genes with detected expression in the filtered counts matrix. Genes ranked by log2 fold change were used as an ordered query. P values were corrected using the g:SCS algorithm to account for multiple comparisons.

### Cell type gene signature enrichment analysis

Single-cell immune cell expression signatures derived from healthy PBMCs^87^ were used for gene set enrichment analysis against the DGE lists of each comparison to infer the cellular composition of the RNA-seq signatures. Enrichments were performed using Fisher’s exact tests and using Bonferroni correction for multiple comparison (p ≤ 0.05).

### Cell type deconvolution with UniCell Deconvolve

We used a deconvolution method that leverages embeddings from pre-trained data using an extensive cell type database using UniCell Deconvole^52^. First, we applied a read-depth normalization with additional TMM normalized factors to the raw RNA-seq counts, followed by log_2_ transformation. We then applied a contextualized prediction with UCDSelect, using a built-in PBMC-specific reference with 30 cell types but ignoring the “native-cell” category. The results were summarized across all samples computing the mean predicted fractions per group (mild, severe) and collection time point (days 0, 3, 7 and 46). Mean estimates were then plotted as mean cell type proportions across all cell predicted types (Figure 3D and Supplementary Figure 3). For each cell type, Mann–Whitney *U* test was performed to compare differences between mild vs. severe patients at each time point. Statistical significance was considered when p ≤ 0.05.

### Olink

We measured cytokines in plasma using Olink multiplex proximity extension assay (PEA) inflammation panel (Olink proteomics: www.olink.com) according to the manufacturer’s instructions. The PEA is a dual-recognition immunoassay, in which two matched antibodies labelled with unique DNA oligonucleotides simultaneously bind to a target protein in solution. This brings the two antibodies into proximity, allowing their DNA oligonucleotides to hybridize, serving as template for a DNA polymerase-dependent extension step. This creates a double-stranded DNA ‘barcode’ that is unique for the specific antigen and quantitatively proportional to the initial concentration of target protein. The hybridization and extension are immediately followed by PCR amplification, and the amplicon is then finally quantified by microfluidic qPCR using Fluidigm BioMark HD system (Fluidigm). Processing and quality assessment of proteomics data were performed using the “Olink NPX manager” software. The data was transformed and normalized to Olink’s NPX value. NPX is a relative protein quantification unit on a log2 scale where a difference of 1 NPX equates to a doubling of protein concentration.

### Fc-binding protein array

The binding of S antigen specific antibodies to human FcγR was analyzed by Luminex technology. S antigens were coupled to Luminex beads (Luminex Corp, TX, USA) by carbodiimide-NHS ester-chemistry with an individual region per antigen. Coupled beads were incubated with diluted plasma sample (1:2,000 for FcγR probing) for two hours at room temperature in 384 well plates (Greiner Bio-One, Germany). Next, unbound antibodies were washed away. For the analysis of FcγR binding PE-Streptavidin (Agilent Technologies, CA, USA) was coupled to recombinant and biotinylated human FcγR2AH/R, FcγR2B, FcγR3AF/R and FcγR3B (Duke Human Vaccine Institute Protein Production Facility). Coupled FcR were used as a secondary probe at a 1:1000 dilution. After one hour incubation, excessive secondary reagent was washed away and the relative antibody concentration per antigen determined on an iQue analyzer (IntelliCyt). Each sample was analyzed in duplicates.

### THP-1 monocyte phagocytosis assay

An assay for measuring antibody-dependent THP-1 monocyte phagocytosis was used as previously described^98^. S antigens were biotinylated using an NHS-Sulfo-LC-LC kit according to the manufacturer’s instruction (Thermo Fisher Scientific). Excessive biotin was removed by size exclusion chromatography using Zeba-Spin desalting columns (7kDa cutoff, Thermo Fisher Scientific). Biotinylated S antigens were coupled to FluoSphere NeutrAvidin beads (Thermo Fisher Scientific) and incubated with 10 μl 1:10 diluted plasma for two hours at 37°C to form immune complexes. THP-1 monocytes (American Type Culture Collection) were added to the beads, incubated for 16 hours at 37°C, washed and fixed with 4% paraformaldehyde. Samples were analyzed on an iQue analyzer (Intellicyt). Each sample was analyzed in duplicates

### Primary neutrophil phagocytosis assay

Phagocytosis score of primary human neutrophils was determined as described before^99^. S antigens were biotinylated and coupled to fluorescent neutravidin beads (Thermo Fisher) and incubated with 1:10 diluted plasma. Primary cells were derived from Ammonium-Chloride-Potassium (ACK) buffer lysed whole blood from healthy donors and incubated with immune complexes for one hour at 37°C. Neutrophils were stained for surface CD66b (BioLegend, Cat.# 305112) expression, fixed with 4% paraformaldehyde, and analyzed on iQue analyzer (IntelliCyt)

### Complement deposition assay

The complement deposition assay was performed as previously described^100^. S antigens were biotinylated and coupled to FluoSphere NeutrAvidin beads (Thermo Fisher Scientific) and incubated with 10 μl 1:10 diluted plasma samples for two hours at 37°C. After non-specific antibodies were washed away, immune-complexes were incubated with guinea pig complement in GVB++ buffer (Sigma-Aldrich) for 20 minutes at 37°C. Complement reaction was stopped with EDTA-containing phosphate-buffered saline (15mM) and C3 deposition on beads was stained with a 1:100 diluted anti-guinea pig C3-FITC antibody (MP Biomedicals, Cat.# 0855385) and analyzed on an iQue analyzer (Intellicyt). Each sample was analyzed in duplicates.

### Statistical analysis

All statistical analyses were performed using GraphPad Prism (v9). Continuous variables were summarized as median with interquartile range (IQR) unless otherwise indicated. For comparisons between two independent groups (mild vs. severe) at a given time point, two-tailed Mann–Whitney U tests were used. Categorical variables were compared using Fisher’s exact test when applicable. Correlation analyses were performed using Spearman’s rank correlation. For RNA-seq differential gene expression, linear modeling was performed using Limma with voom transformation; empirical Bayes moderated statistics were used, and p values were corrected for multiple testing using the Benjamini–Hochberg method (adjusted p ≤ 0.05).

For cell-type gene signature enrichment, Fisher’s exact tests were applied with Bonferroni correction for multiple comparisons (p ≤ 0.05). For Gene Ontology enrichment analyses, multiple testing correction was performed using the g:SCS algorithm. For Olink proteomics differential analyses, p values were adjusted for multiple testing using the Benjamini–Hochberg method as indicated. Principal component analyses (PCA) were performed on scaled features, and 95% confidence ellipses were used for visualization. Statistical significance was defined as p ≤ 0.05 unless stated otherwise.

## Data Availability

All data are available in the manuscript or the supplementary materials.

## ACKNOWLEDGEMENTS

We thank the BACO cohort and their families. We also thank Richard Cadagan for excellent technical assistance and Jenny S. Maron for technical performance and support on this project. This work was partly funded by CRIPT (Center for Research on Influenza Pathogenesis and Transmission) and NIAID-funded Center of Excellence for Influenza Research and Response (CEIRR, contract # 75N93021C00014) to A.G.-S and T.A. and the Systems Biology Lens (SYBIL, # U19AI135972) and the Viral Immunity and Vaccination (VIVA) Human Immunology Project Consortium (HIPC) (# U19AI168631) to A.G.-S. T.A. was also funded by the American Lung Association: ALA grant # COVID-1034091.

## CONTRIBUTIONS

T.A. and A.G.-S. conceived and designed the study. A.E, A.S.G-R and T.A analyzed all the data and prepared figures. A.E performed ELISAs assays. S.A. generated stable A549 HexaPro Spike protein cell line and performed ADCC/P assays. E.B. performed intermediate analysis on Fc responses and Olink proteomics. J.S.M. performed binding and functional Fc-responses. A.R.-F. performed ELISA for SARS-CoV-2 full-length S protein. A.R., G.A.A., and J.C. collected samples and clinical data. M.A.A. and V.D.N performed RNA extractions and library prep. H.v.B. and G.A. provided expertise. T.A. and A.G.-S. supervised the study. A.E. and T.A. wrote the manuscript. All the authors reviewed and edited the manuscript.

## DATA AVAILABILITY

All data are available in the manuscript or the supplementary materials.

