## Supplementary Figure 1 for "Early Fc-effector antibody signatures impact COVID-19 disease trajectory"

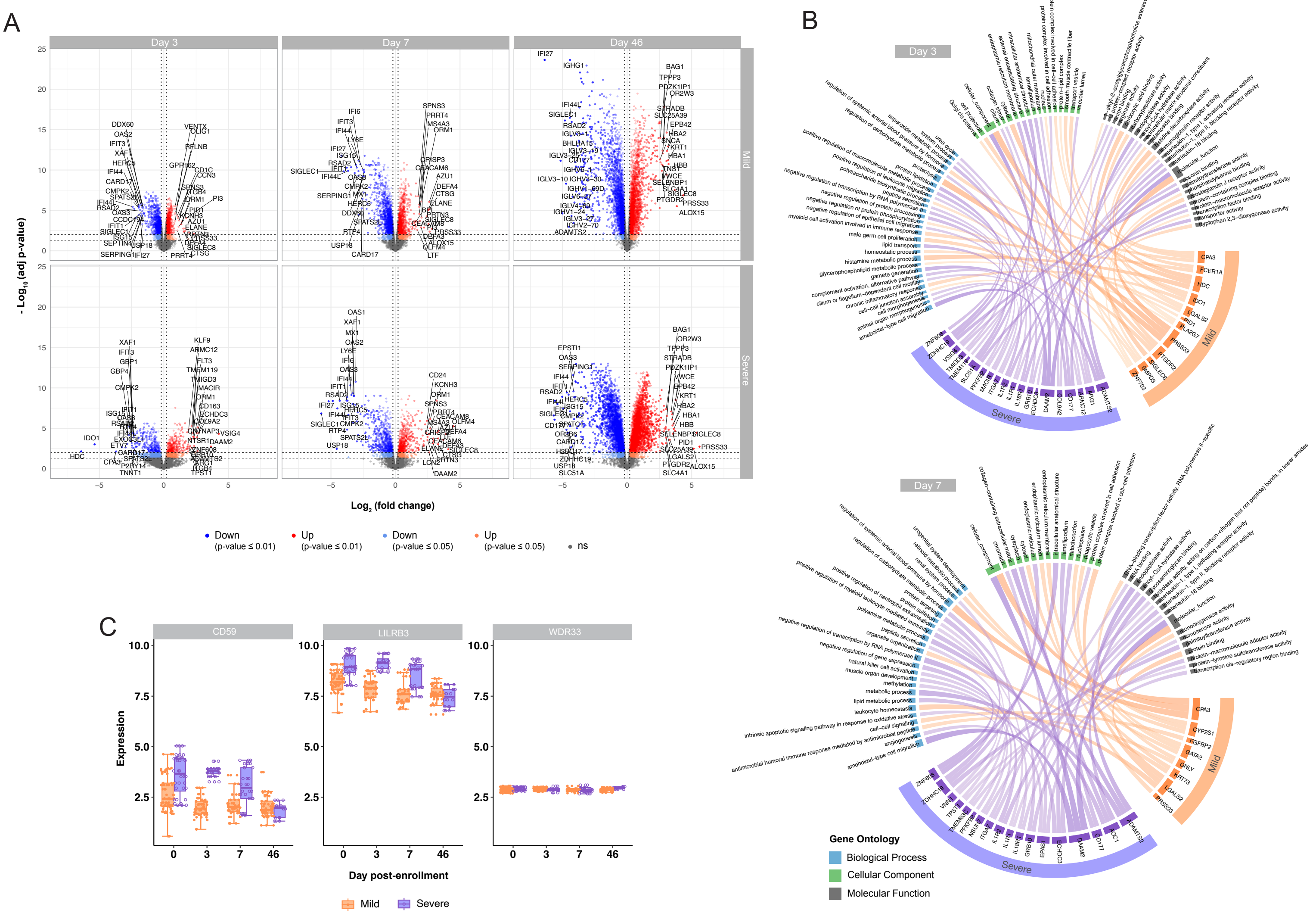


**Supplementary Figure 1. Differential gene expression and pathway enrichment in mild and severe COVID-19 patients. A)** Volcano plots of upregulated (red) and downregulated (blue) genes in mild and severe COVID-19 patients on days 0, 3, and 7 post-recruitment. Expression differences > 1.5-fold change are shown (log2 fold change ≤ 0.5, vertical dotted lines). Darker colors show genes with an adjusted p-value ≤ 0.01, while lighter colors show adjusted p-value ≤ 0.05. The names of the top 20 genes are shown. **B)** Chord diagram of Top 20 upregulated genes and their assigned GO pathways in mild and severe patients at day 3 and 7 post-enrollment. **C)** Box-and-whisker diagrams showing absolute expression of LILRB3 gene in mild and severe COVID-19 patients from the BACO cohort. CD59 and WDR33 were included as controls.
