## Supplementary Figure 2 for "Early Fc-effector antibody signatures impact COVID-19 disease trajectory"

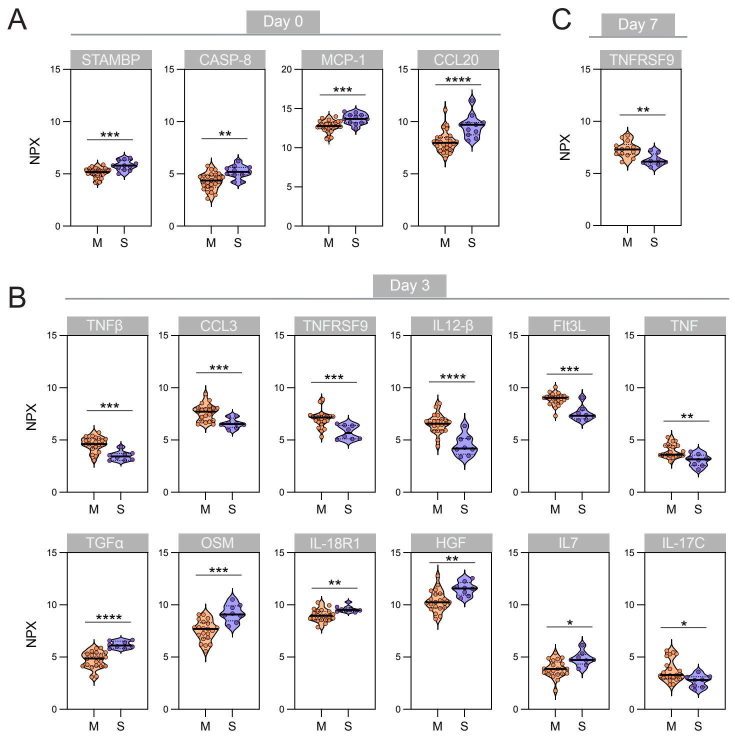


**Supplementary Figure 2. Cytokine expression in mild and severe COVID-19 patients over time.** Violin plots showing differentially expressed cytokines in mild and severe COVID-19 patients on days 0 **(A)**, 3 **(B)**, and 7 **(C)** post-recruitment. Mann-Whitney U test was performed to compare differences between mild vs. severe patients at each time point. Statistical significance was considered when $p \leq 0.05$ (*$p < 0.05$, **$p < 0.01$, ***$p < 0.001$, ****$p < 0.0001$).
