## Supplementary Figure 3 for "Early Fc-effector antibody signatures impact COVID-19 disease trajectory"

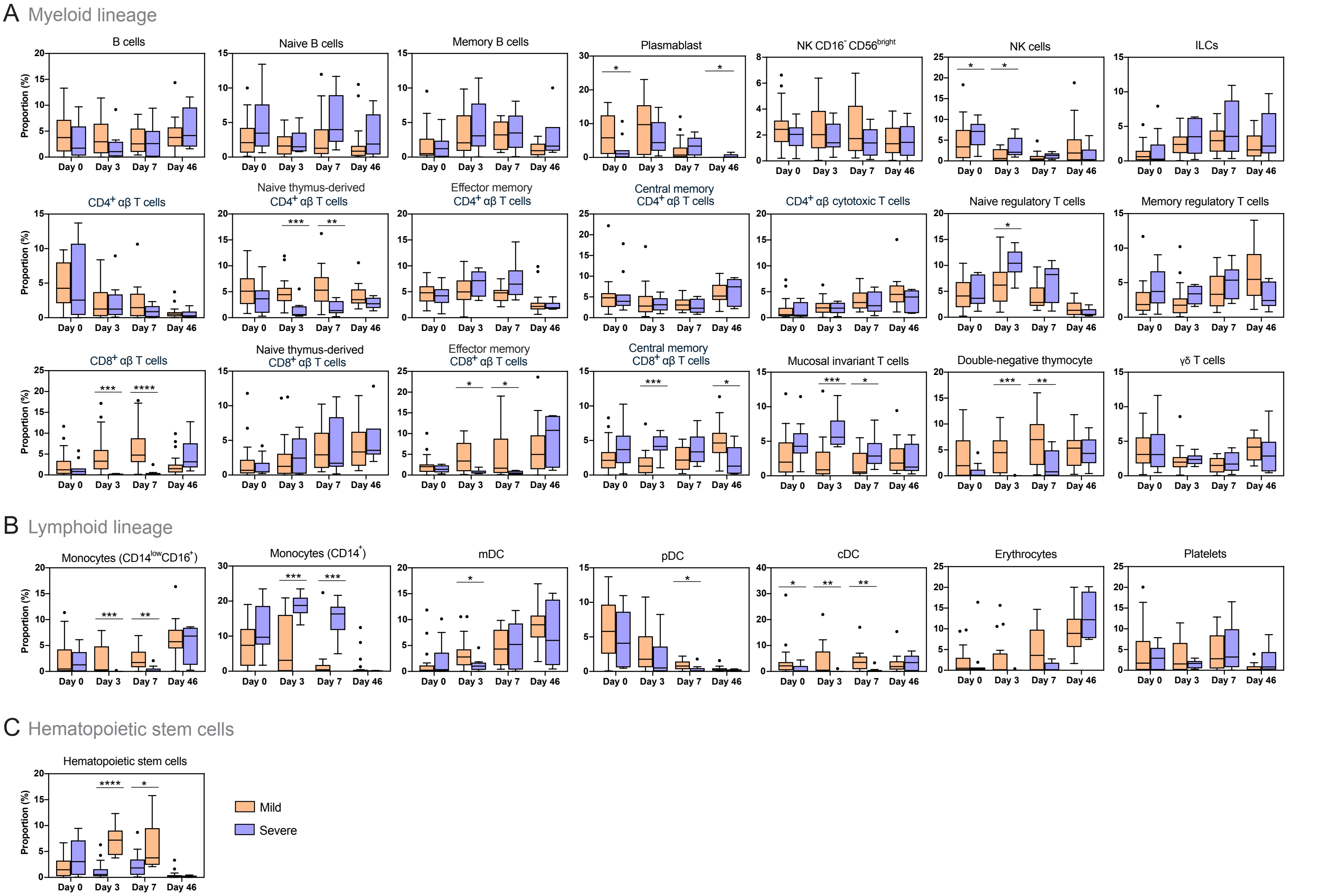


**Supplementary Figure 3. Immune cell lineage composition inferred by the UCD method in mild versus severe COVID-19 patients.** Box-and-whisker plots showing the relative abundance of immune cell types from **(A)** the myeloid lineage, **(B)** lymphoid lineage and **(C)** hematopoietic stem cells in mild and severe COVID-19 patients. Box indicates interquartile range (IQR, Q1-Q3), with horizontal line showing the median and vertical lines indicating minimum and maximum. Mann-Whitney U test was performed to compare differences between mild vs. severe patients at each time point. Statistical significance was considered when $p \leq 0.05$ (*$p < 0.05$, **$p < 0.01$, ***$p < 0.001$, ****$p < 0.0001$).
