## Supplementary Figure 4 for "Early Fc-effector antibody signatures impact COVID-19 disease trajectory"

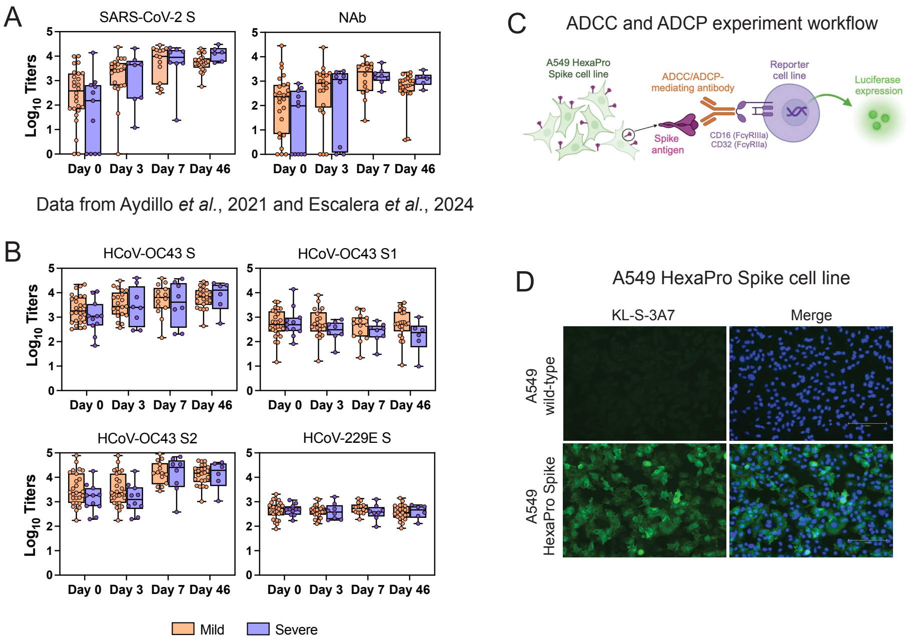


**Supplementary Figure 4. Humoral immunity to SARS-CoV-2 and seasonal HCoV S antigens in the BACO Cohort.** Box-and-whisker plots showing **(A)** binding antibodies titers against SARS-CoV-2 full-length S protein and neutralizing antibodies against authentic SARS-CoV-2 Wuhan-like virus **(B)** the binding antibodies titers against HCoV-OC43 full-length S, S1 and S2 antigens and HCoV-229E full-length S in mild and severe COVID-19 patients. Box indicates interquartile range (IQR, Q1-Q3), with horizontal lines showing the median and vertical lines indicating minimum and maximum. Mann-Whitney U test was performed to compare differences between mild vs. severe patients at each time point. Statistical significance was considered when $p \leq 0.05$. **(C)** Overview of the ADCC and ADCP assay workflow. D) Validation of A549 HexaPro Spike cell line. Spike protein was stained with primary anti-body KL-S-3A788. DAPI (4¢,6-dia-midino-2-phenylindole) was used to visualize the nucleus.
