## Supplementary Figure 5 for "Early Fc-effector antibody signatures impact COVID-19 disease trajectory"

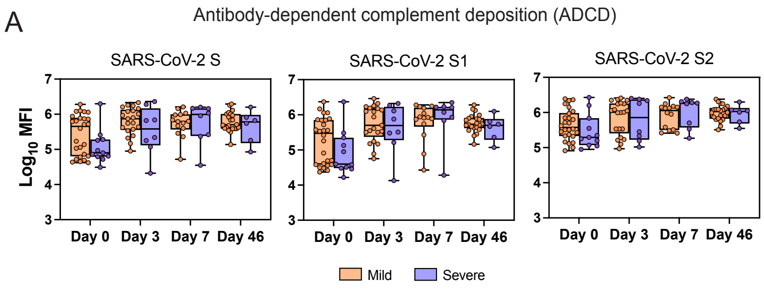


**Supplementary Figure 5. Antibody-dependent complement deposition responses to SARS-CoV-2 S antigens in mild and severe COVID-19. A)** Box-and-whisker plots showing antibody-dependent complement deposition (ADCD) responses against SARS-CoV-2 full-length S, S1 and S2 antigens in mild and severe COVID-19 patients. Box indicates interquartile range (IQR, Q1-Q3), with horizontal lines showing the median and vertical lines indicating minimum and maximum. Mann-Whitney U test was performed to compare differences between mild vs. severe patients at each time point. Statistical significance was considered when $p \leq 0.05$.
